# Adoption of Community Adolescent Treatment Support for HIV Care Retention in Greater Accra Region, Ghana

**DOI:** 10.1101/2024.11.27.24318064

**Authors:** Margor A. Green-Harris, Thembikile Nyasulu, Jordanne Futu Ching, Robert Kogi, Emmanuel Asampong, Phyllis Dako-Gyeke

## Abstract

Despite the introduction of efficacious antiretroviral therapy, HIV and AIDS continue to pose a threat to global public health. Community adolescent treatment support programs have shown promising signs of improving adherence retention to care which in turn improves HIV-related health outcomes in this population. However, little is known about the adoption of this program among adolescents in Ghana. This study sought to assess the adoption of community adolescent treatment support among adolescents living with HIV in Tema General Hospital and LEKMA Hospital in the Greater Accra Region, Ghana.

A qualitative phenomenological design was carried out at the Tema General Hospital and LEKMA Hospital. Adolescents living with HIV aged 18 to 24 years old who were actively participating in the Community Adolescents Treatment Support (CATS) services were recruited in this study. In addition, healthcare workers involved in the CATS strategy since its inception were recruited. A total of sixteen (16) key informant and in-depth interviews were conducted. The data was transcribed verbatim and imported to NVivo 14.0 software for thematic analysis.

It was found that CATS serves as a crucial support system, strengthening peer connections and providing flexible medication pickup. Improved medication adherence, better retention in care, and positive viral load outcomes were evident among participants. However, communication issues and participants consistency pose challenges. Support group meetings were highly valued by participants. Mobile health services were widely accepted for maintaining connectivity and support. Home visit services faced reluctance due to privacy concerns, and the wider community was perceived as unwelcoming. While challenges persist, including recruitment difficulties and financial support, there is a strong commitment to improving the program’s effectiveness and scalability. Communication strategies, increasing funding support, and intensifying community education and sensitization efforts should be strengthened to reduce stigma and foster broader community inclusivity.

## Introduction

Despite global efforts to curb Human Immunodeficiency Virus (HIV) pandemic, it remains a threat to public health. About 1.5 million new HIV cases were recorded in 2021, which is three (3) times more than global targets [1]. Young adults and adolescents have stood out among persons living with HIV because they represent the only category where HIV-associated mortality is on the rise despite the inception of highly active antiretroviral therapy [2]. HIV is among the five top causes of death in the aged bracket 10-19 [3].

Even though there is limited data on treatment coverage among adolescents, access to and uptake of antiretrovirals (ARV) are often reported to be lower in adolescents [3,4]. Adolescents living with HIV are at a higher risk of loss to follow-up both before ARV initiation and after initiation, with pregnant adolescents living with HIV and adolescents’ key populations, being particularly vulnerable [5].

Addressing the specific yet diversified needs of adolescents living with HIV to improve their HIV-related outcomes requires an all-inclusive approach. Community Adolescent Treatment Supporters is a peer-led model for supporting adolescents and youths living with HIV [6]. This strategy revolves around young people living with HIV (ages 18-24) who are to support 0 to 24-year-olds living with HIV to initiate and remain on treatment [6].

From 2005 to 2017, the global figure of AIDS-related deaths decreased by 48%. However, it increased by 50% among all adolescents and young people [7]. In 2020, Ghana recorded 3,683 and 5,211 new infections in the aged bracket 10-14 and 15-24 [8]. One of the most potent forces backing the high morbidity and mortality rates among adolescents living with HIV is suboptimal adherence to ARV the [9].

A study done in Zimbabwe showed evidence of how adolescents living with HIV using community adolescent treatment support have improved in linkage, ARV adherence, and psychophysical well-being [10]. Another study conducted in Ghana showed that stigma and discrimination remain a barrier to HIV care [11].

A feasibility and acceptability study in Kumasi, Ghana, revealed that an in-clinic peer group is a safe space for connecting adolescents living with HIV [12]. It also suggested that additional work needs to be done to endorse further in-clinic peer groups and their impact on their commitment to care and treatment adherence [12]. These groups can help adolescents living with HIV deal with the emotions around the disease, provide support for handling the daily challenges of disease management and provide a stage to acquire skills in disclosure [13]. Another study in Kumasi showed that adolescents living with HIV have low self-esteem and recommended social support as an intervention [14]. This study therefore explored the adaptability of community adolescent treatment supporters among adolescents living with HIV in Greater Accra region, Ghana.

## Materials and Methods

### Study Design

This study used a hospital-based qualitative phenomenology study design. Maximum variation and convenience sampling were used. This design enabled in understanding the lived experiences and perspective of participants [15,16] on the community adolescent treatment support strategy. A maximum variation technique was used to capture a broad spectrum of perspectives, experiences, and characteristics within the study population [17]. Convenience sampling was used to recruit healthcare workers for the study based on their availability and accessibility.

### Study Area

The study was conducted in two selected hospitals within Greater Accra (Ledzokuku Krowor Municipal Hospital and Tema General Hospital). These hospitals were selected as they have actively implemented community adolescent treatment support programs. The Ledzokuku Krowor Municipal Assembly (LEKMA) Hospital is a government-owned health facility at Teshie. It serves as the municipal hospital for populaces within the Ledzokuku Krowor Municipal area and beyond. The Hospital’s ART services run from Mondays to Fridays, from 8 am to 2 pm. Wednesdays and Thursdays are designated for adolescent services, and on these days, their clinical staff and peer support team are consistently present. Additionally, the final Saturday of each month is specifically set aside to address the needs of clients actively participating in the community adolescent treatment support group. On this designated day, the supervisor, peer supporter, and clients convene for their planned meetings and interactions.

The Tema General Hospital on the other hand, is also a government–owned health facility in the Tema metropolis. Being the largest public health facility in Tema metropolis, it caters to the health needs of the people of Tema metropolis and its satellite towns and villages. They run a daily ART clinic. Every Friday is dedicated exclusively to adolescents, during which they ensure that adolescent-friendly services are readily accessible, the clinical staff members are available, and peer supporters are present for duty. The community adolescent treatment support group sessions are scheduled to take place on Fridays and occur once every three months. On these designated Fridays, the supervisor, peer supporter, and clients come together for their scheduled meetings and interactions.

### Study Population

The target population for this study are three sets of participants, adolescents living with HIV working as community adolescent treatment support (CATS), adolescents living with HIV receiving care from the treatment support (clients) and the Healthcare Workers. To promote gender sensitivity, the researchers aimed to achieve a balanced representation of both male and female participants, aged between 18 and 24, among the clients and their peer supporters involved in the study. However, achieving this gender balance proved challenging as there was a lack of available male peersupporters in the facilities involved for inclusion in the study.

### Inclusion and Exclusion Criteria

Eligible for participation in this study include adolescents and youth aged 18-24 years living with HIV who are either clients or peer supporters in the Community Adolescent Treatment Support Group. Participants were enrolled in either LEKMA Hospital or Tema General Hospital CATS services and must be a participant for at least a period of one (1) year. Participants were fluent in either English language or a local language spoken by the research team. Healthcare Workers involved healthcare workers in the Community Adolescent Treatment strategy since its inception that were willing to participate were also be included in the study.

Those who meet the eligibility criteria but were not part of the study included adolescents and youth aged 18-24 years that were critically ill at the time of the study. Individuals who required specialized care or treatment that was outside the scope of the study. Sampling

### Sample Size Determination

A total of 16 participants were recruited for the study, selected based on their availability and willingness to participate. A total of 3 Key informant interviews and 13 in-depth interviews were conducted 3 with the peer supporters and 13 with the clients. The number of participants aligns with the standard number of participants needed to reach data saturation in a phenomenological study [18,19]. The key informant interview aimed at getting first-hand information from the health workers, and the in-depth interview elicited the personal experiences and perspectives of the Community Adolescent Treatment Support clients and peer supporters. However, the interviews were stopped when a point saturation is attained [20,21]. Saturation was considered to have occurred when the participants added later did not significantly contribute new information [22].

### Sampling Technique

A purposeful sampling technique was to identify and select study participants based on their experience with the community adolescent treatment support and knowledge about the intervention [19,23]. The study employed the maximum variation sampling technique to carefully select participants from ART clinics. This technique aims to achieve a diverse and varied sample by deliberately including individuals with a wide range of characteristics such as age, gender, socio-economic status, educational background, and other relevant factors [17].

By purposefully incorporating participants with diverse backgrounds and perspectives, the study captured a comprehensive understanding of the topic at hand. This approach contributed to a richer and more nuanced exploration of different viewpoints, contexts, and experiences, ultimately enhancing the depth and relevance of the research findings [17,24]. Eligible participants were intentionally selected based on peculiar characteristic [20]. Permission was sought from the health directorate with the help of the introductory letter from the school of public health and the ethical clearance. The supervisors in charge of the service helped to identify peer supporters and clients using the community adolescent treatment supporters using the hospital records. Upon agreeing to participate in the research, the study’s purpose and the consent form’s content was explained to the participants in detail. Eligible and interested participants were given a consent form to sign before participation. Healthcare workers involved in the Community Treatment Support intervention were sampled using a convenience sampling technique based on their willingness and availability [17,20].

### Data Collection and Tool

The semi-structured interview guide was pretested at Weija Gbawe Hospital, where the intervention has been rolled out. The data was reviewed to suit the study population. The interview guide was used to conduct the interviews to get the required information from the participants. It is an effective qualitative tool for accessing profound thoughts and personal experiences surrounding the program [25]. The interview guide was divided into various sections covering the perceived effectiveness of community adolescent treatment support for linkage to HIV care and follow-up among adolescents, the perception of adolescents towards community adolescent treatment support, the community factors that influence the implementation of Community Adolescent Treatment Support, the healthcare worker’s perceptive on community adolescent treatment support and constructs from the conceptual framework [25]. The interview guide for the healthcare workers looked at the experience and challenges in service delivery. Other tools used in the study include digital recorders, pens, pencils, and field notebooks to record and document information and codebook to generate themes [18]. The recruitment period and data collection of the study started on 1st August 2023 to 28th August 2023.

### Data Collection Technique

Key informant interviews (KIIs) were conducted with healthcare workers and In-depth Interviews (IDIs) were conducted with community adolescent treatment support clients and peer supporters respectively. The Key Informant Interview was conducted in the respective health facility with the supervisors involved in the Community Adolescent Treatment Support strategy that were available and willing to participate at the time of the study. The In-depth Interviews was with adolescents living with HIV who are either clients or work as peer support [26]. They comprised of community adolescent treatment supporters that meet the eligibility criteria and are willing to participate at their convenience [27]. Each session lasted for an average of 15 to 20 minutes.

### Data Analysis

The recording was carefully listened to thrice before being typed verbatim into Microsoft Word version 19. The written transcript was imported to NVivo 14.0 software for analysis after it was proofread, inductive and deductive framework was used in developing the codebook, coding scripts, and developing themes [28,29]. The conceptual framework and objectives of the study interview guide served as a guide for the preliminary development of the codebook [28, 29]. The codebook was revised to include the emerging themes from the collected data [29,30]. The codebook was discussed with and accepted by both the principal investigator and the supervisor.

### Quality Control

A two-day training was organized for the research assistants. The training aims to explain the study’sobjectives, the importance of obtaining consent and assent and maintaining confidentiality. The team pretested tools to guarantee the accuracy and precision of data. The data collection tool was modified. The transcribed audio interviews were double-checked with the written notes avoid to missing out on critical information. Transcripts were shared with participants to solicit their feedback, validation, and clarification on the study findings. Simultaneously, the remaining transcripts were distributed among the research team members. After each data collection session, the research team conducted debriefing sessions to discuss and analyze the collected data.

### Ethical Consideration

Ethical clearance was sought from the Ghana Health Service Ethics Review Committee with reference number: **GHS-ERC:070/05/23**. Permission was sought from the Greater Accra Regional Health Directorate and from the management of the hospitals and the respective heads of department of each health facility.

The participants signed a participant information sheet with an informed consent form before the data collection and interview. Participants were also allowed to decide not to participate, ask questions if they thought it was uncomfortable, or leave the study whenever they wanted. The discussion and interviews were conducted in a private setting for privacy purposes. All data received was protected and was not accessible to individuals who were not part of the team. Participants were made aware that the findings of this would be published in peer-reviewed journals. The findings and recommendations from this study were shared with the study participants and the hospitals where the study took place.

## Results, Discussion, Conclusion

### Results

#### Socio-demographic Characteristics of Respondents

In this study, a total of 16 participants were involved. The adolescent participants hereby referred to as clients ranged in age from 18 to 24 years. Three treatment supporters, commonly known as CATS, who were individuals living with HIV and serve as peer supporters within their respective healthcare facilities. The age range for these CATS was between 23 and 24 years. Three healthcare workers, aged 33-49 years, consisting of one technical supervisor responsible for overseeing community treatment support at the Christian Health Association of Ghana (CHAG), and two supervisors from the respective healthcare facilities. In terms of gender distribution, the clients had an equal number of males and females, whereas all the Peer Supporters were females. Out of the three supervisors, two were females, constituting approximately 66.7% of the supervisory team, while the technical supervisor from CHAG was a male. A greater proportion of adolescents (69.2%) of them had attained basic education level whereas all the supervisors (100%) obtained tertiary education.

In addition, majority of the Clients and Peer Supporters (84.6%) were Christians, whilst all the supervisors were Christians. A little above half (53.8%) of the Clients and Peer Supporters were formally employed and a greater proportion (61.5%) of the respondents were single. All healthcare workers were formally employed whilst 2 out of 3 were married.

#### Effectiveness of community adolescent treatment support in facilitating HIV care linkage and follow-up among adolescents

It was reported that the program’s method of attracting clients is simple. Healthcare workers achieved this by engaging clients at the standard ARV clinic, presenting them with information about the program and its offerings. Notably, there was a greater willingness among girls to join the program, surpassing the interest from boys. Many clients find it appealing, highlighting its effectiveness, as demonstrated by the excerpts below.

> *“I was informed by one of the senior nurses there then yeah! a lady took me through then added me, then ahh I decided [to be part of it”* (In-depth Interview Participant, Client, LEKMA).

> *“We sell the idea to them and their caregivers and almost all of them over 70-80 percent are interested”* (Key Informant Interview Participant, Healthcare Worker, TEMA).

> *“We recruit them through our respective agents the ARV supervisors.”* (Key Informant Interview Participant, Healthcare Worker, CHAG).

The motivations driving adolescents to engage with this program resembled individuals’ needs and perspectives. One salient observation was that CATS offers a secure and welcoming environment for adolescents to engage, acquire knowledge about HIV, and establish meaningful connections. Some adolescents were enticed by the knowledge they will gain. The excerpts below illustrate these findings.

> *“I decided to join to learn the pros and cons of this virus or this infection and to get more insight about what to do or what not to do and just to be a bit encouraged. When I get some mind-boggling questions too, I [did] ask them for clarification.”* (In-depth Interview Participant Client, LEKMA)

> “*I just wanted to link up, so [that] I’ll know more about this (HIV)”* (In-depth Interview Participant Client, TEMA)

Also, being part of the CATS program was driven by the longing for emotional support and the comfort of belonging among peers who share similar life experiences. There were also those who attended out of a sense of duty or perhaps a more subdued interest, contributing to the program’s motivations. In addition, it serves as a crucial support system, specifically designed to address the unique emotional, social, and psychological needs of adolescents living with HIV as shown in the following excerpts.

> *“The support group provides the opportunity for you to share your life experiences, ask questions about life, share the experiences of what you go through daily, how you feel about this whole thing (HIV), like Emm! Just trying to normalize everything regardless of you having the virus. … So, it gives that scenario or creates that scenario for you to talk more about your life experiences.”* (In-depth Interview Participant, Client, LEKMA).

> *“I’m supposed to be there to encourage that person or to alert that person that is not a death sentence,uh, virus or sickness.”* (In-depth Interview Participant, Peer Supporter, TEMA).

Peer supporters maintain a comprehensive list of their clients and regularly make proactive phone calls to ensure their well-being, remind them of upcoming appointments, and provide valuable support. Furthermore, clients benefit from the flexibility of requesting drug pickups beyond the standard clinic hours or location, underscoring the program’s accessibility and adaptability, as exemplified in the excerpts below.

> *“They call to check on me and deliver my drugs”* (In-depth Interview Participant, Client, TEMA)

> *“Then I will take ok, you can come on Saturday to take it, but if it’s not closer to the support group. I would just be like Oh, come meet me somewhere and take your medicine.*” (In-depth Interview Participant, Peer Supporter, LEKMA)

> “*The CATS have the contact of most of her peers, so whoever is supposed to come for a refill on a particular day and they are not able to come sometimes they call, oh am supposed to [be here] this week but I can’t come, can you pick it up so I pick it during meeting days, so we pick it up and then we keep it and then on meeting days they come and pick it up.”* (Key Informant Participant, Healthcare Worker, LEKMA)

Clients have consistently reported noticeable improvements in their adherence and retention into care,which is substantiated by better viral load outcomes and a reduced need for hospital visits. These sentiments have been reiterated by both Peer Supporters and supervisors as shown in the following excerpts.

> *“The last time that we had our support group one of my colleagues shared her experience. So, from that day it motivated our clients to take the medicine, the last time that one [of] our clients came to the facility to take her medicine, she was looking good, but when she wasn’t taking it, …she was [having] rashes all over her body, like she was becoming slim”* (In-depth Interview Participant, Peer Supporter, TEMA)

> *“So I can say that it is improving adherence now because they know why they are supposed to be taking their medications and then they know”* (Key Informant Interview Participant, Healthcare Worker, LEKMA)

> *“So, before the support group formation we [had] some of them defaulting on treatment. It is difficult staying loyal to a thing such as this because for most of them they were diagnosed when they were children and they [have] been on medication for a long time, so when they become adolescent the teenage day especially when they are in the boarding schools most of them default…. doing well on medication,having fewer hospital visits because of illness and all that they started taking their drugs and [their] parents were happy that at least their children and wards got to know that with adherence they could live normal healthy lives”* (Key Informant Interview Participant, Healthcare Worker, TEMA).

While widely embraced for its invaluable support to adolescents living with HIV, the CATS program is confronted with specific challenges in its implementation. Notably, it was revealed that the program grapples with issues related to communication, financial barriers, and participant consistency as depicted in the following excerpts.

> *“At times after my appointment that would be where they will be telling me that they had a meeting [and] they forgot to call me”* (In-depth Interview Participant, Client, TEMA)

> *“In order to improve our support group, … like financial, unless we support them with their TnT”* (In-Depth Interview Participant, Peer Supporter, TEMA)

> *“Most of my clients, …when you call them to come for the meeting they’ll be like, Ok, I’ll come but when it’s time they’ll be like Oh, I’m sorry, I’m going to work, I’m busy.”* (In-depth Interview Participant, Peer Supporter, LEKMA)

#### Acceptance of community adolescent treatment support by adolescents

The prevailing sentiment among the participants was the indispensable value of the companionship provided by the support group; a sentiment strongly echoed by all the participants. In the quest to provide comprehensive healthcare and support for adolescents, the program utilizes various strategies and services. All the adolescents involved in the program enthusiastically embraced it because it provided them with the reassurance that they are not alone. They found that the group meetings created a welcoming and secured space where they could freely express themselves and be their true selves.

> *“It’s fun and we are all the same age group living with HIV so it’s fun, hearing of things like living a positive life, hearing how to cope with HIV and all.”* (In-Depth Interview Participant, Client, LEKMA).

> *“I didn’t want anyone to know I had HIV. I was so scared that people would judge me, and I felt like I’d be alone but when I came to the group meetings, I realized I’m not alone. There are others like me,and they support me. It’s a safe space where I can be myself.” (In-Depth Interview Participant, Client,TEMA)*

The integration of technology and healthcare has emerged as a powerful tool for maintaining connectivity and extending support to adolescents within groups. Mhealth, encompassing the use of SMS and phone calls to facilitate communication between CATS and their clients, has become a cornerstone of the services. This service provides many opportunities for staying connected with clients, offering a means of ongoing care and support. Mhealth services have not only enabled the peer supporters to reach out promptly but have also allowed adolescents to access information, ask questions, and receive guidance at their convenience. It is the only service that was accepted by all the participants. Notably, this service garnered unanimous acceptance among all participants as shown in the following excerpt.

> *“It has made it so much easier to reach out when I need support, I can just drop a message, and they will get back to me. It feels more private and less intimidating.”* (In-Depth Interview Participant, Client, LEKMA).

> *“You’ll be down once a while they will call you as if they know that maybe you are down, and they will just encourage you and they will give you hope.”* (In-depth Interview Participant, Client, LEKMA).

> *“It makes me feel like someone is thinking about me, like they care for me.” (In-depth Interview Participant, Client, TEMA)*.

The convenience provided by this service has led to some female clients expressing a desire for additional features, with a few suggesting that a WhatsApp platform could serve as a virtual meeting space as demonstrated by the following excerpt.

> *“They kindly create a group… So, it would be like even outside meeting, you will still have an area where you can connect”* (In-Depth Interview Participant, Client, TEMA).

> *“I find WhatsApp convenient. You can quickly send a message or even voice notes. It’s a good way to keep in touch and get support when you need it. Plus, it doesn’t require a lot of data, which is important for some of us.” (In-depth Interview Participant, Client, LEKMA)*.

Despite its merits, this approach encounters challenges, as not everyone possesses a smartphone, and some may be restricted from using phones during school hours, and privacy concerns persist as shown by the following excerpt.

> *“I tried creating that but not most of the clients are on WhatsApp”* (In-Depth Interview Participant, Peer Supporter, LEKMA).

> *“I’m a bit hesitant about joining a WhatsApp group. I’m not comfortable with the idea of everyone in the group having my phone number. I prefer more private ways to connect with the program.” (In-depth Interview Participant, Client, TEMA). “Some of my clients are in boarding schools and are not allowed to use phones, which can result in them feeling excluded or missing important information.” (In-depth Interview Participant, Peer Supporter, TEMA)*.

Despite the widespread acceptance, some clients persisted in clinging to conventional methods (Standard ARV clinic), while others believed that both approaches are acceptable as can be seen in the extracts below.

> *“I started with the support group, so, I will not know about the other one (Standard ARV)”* (In-Depth Interview Participant, Client, TEMA).

> *“Both of them are good in their own ways because some of them you are able on some days on those different groups you are able to express yourself very well because where there are a lot of people you can’t express yourself but where sometimes we are few you are able to ask some questions, you are able to express yourself. So, I will say both are good in their own ways but depending on the turnout.”* (In-Depth Interview Participant, Client, LEKMA).

At the onset, most adolescents grappled with feelings of embarrassment and a strong inclination toward loneliness. However, as the program advanced, they underwent a remarkable transformation, emerging as confident and independent individuals. This signifies the program’s significant positive influence on the self-sufficiency of clients as demonstrated by the excerpts below.

> *“I am not even comfortable with seeing my other colleagues or people with the same status as me. Sometimes it’s a bit embarrassing honestly”* (In-Depth Interview Participant, Client, LEKMA).

> *“…they normally kept to themselves before the program began. Initially when they come for refills, they are in the background they weren’t even talking. [Sometimes] it’s their parents [who] were coming forward, but since the group started, most of them are now coming on their own to come and pick their medication. Even the meetings we started they were coming with their parents some of them were coming with their caregivers so they will sit outside and wait for us then when we are done, they go but now am sure Saturday you didn’t meet any one with the caregiver.”* (Key Informant Interview Participant, Healthcare Worker, Tema).

While most participants feel comfortable and eager to foster connections, a few grappled with feelings of discomfort as illustrated by the following excerpts.

> *“I feel that is too open even it’s like selected, it’s like those in the same situation.’* (In-Depth Interview Participant, Client, LEKMA)

> “*I’m uncomfortable asking questions openly because some group members are younger than me and are not married.” (In-Depth Interview Participant, Client, TEMA)*

Home visit services are a vital component of the program, providing an opportunity for personalized care and support. However, almost all the participants shy away from this service. The common thread found centered on issues of privacy and stigma. Most of them reside in rented apartments, and concerns about the disclosure of their status to neighbours have led to hesitation about receiving home visits as shown by the following excerpts.

> “I don’t want people to know my business. I live in a rented apartment; you never know who might see them or hear them. It’s just not worth the risk kraa.” *(In-depth Interview Participant, Client, TEMA)*.

> *“When I [called] to arrange a home visit, most, if not all of my clients declined, and I understand them” (In-depth Interview Participant, Peer Supporter, LEKMA)*.

> *‘These refusals are often rooted in a delicate balance of concerns revolving around privacy and stigma.’ (Key Informant Interview Participant, Healthcare Worker, TEMA)*.

#### Community factors that influence the implementation of Community Adolescent Treatment Support

The prevailing viewpoint among most participants is that due to stigma and discrimination, many of them withdraw from social interactions, as they commonly view the community as inhospitable, leading to their discouragement. As such, amidst the support found within the group, there exists an absolute contrast in the way they feel the wider society perceives the condition. As these young voices revealed, discrimination emerges as an intimidating adversary, casting a long and shadowy pall over the lives of those affected as illustrated by the extracts.

> *“The community won’t accept you that’s why I don’t tell anybody about it even my mum.”* (In-depth Participant, Client, LEKMA)

> *“When am having normal conversations with my friend, how they perceive or how they react when they hear it (HIV) is so bad”* (In-Depth Interview Participant, Client, TEMA)

> *“Due to stigma, discrimination and all of that, the young ones are not so willing to come out and then you know, for you to interact them.” (Key Informant Interview Participant, Healthcare Worker, CHAG)*

Almost all the participants concurred that a substantial segment of society remains enveloped in ignorance, as evidenced in the excerpts below.

> *“They are afraid to get close to the person who is affected with the thing (HIV)” (In-Depth Interview Participant, Client, TEMA)*

> *“Their knowledge on modes of transmission and all of that was very very low… they have serious misconceptions about the whole issue regarding you know HIV.” (Key Informant Interview Participant, Healthcare Worker CHAG)*

Most participants share the view that the community is unwelcoming towards individuals living with the virus [HIV] as illustrated by the extracts.

> *“They are not supportive at all.”* (In-depth Interview Participant, Peer Support, TEMA)

> *“People will not see it as just a sickness but will isolate themselves from you, it’s bad*.” (In-depth Interview Participant, Client, TEMA).

The healthcare workers hold diverse perspectives on the involvement of the community, aiming to dispel the shroud of misunderstanding and stigma as demonstrated by the extracts.

> *“We have been trying to [do] periodic engagement with the… community leaders’ religious leaders and things.”* (Key Informant Interview Participant, Healthcare Worker, CHAG)

> *“We haven’t gotten there yet because of stigma; I wonder how many of those adolescents’ parents will allow people know what CATS is about).”* (Key Informant Interview Participant, Healthcare Worker, LEKMA)

> *“So, for now we have not involved the community because HIV thing is quite sensitive.”* (Key Informant Interview Participant, Healthcare Worker, TEMA)

In an earnest effort to fight HIV/AIDS-related stigma and discrimination, healthcare workers are actively involved in initiatives aimed at eliminating both external and self-stigma. This is illustrated by the excerpts below.

> *“We sought to involve religious and community leaders in our efforts because their influential roles within the community can be instrumental in erasing the stigma associated with this issue”* (Key Informant Interview Participant, Healthcare Worker, CHAG)

> *“We conduct counselling sessions to assist them in overcoming self-stigma.”* (Key Informant Interview Participant, Healthcare Worker, TEMA)

#### Stakeholder’s Perceptive on Community Adolescent Treatment Support

Healthcare professionals held the view that the program is effectively addressing the distinctive psychosocial requirements of adolescents, an issue frequently neglected by the current healthcare system. Additionally, they emphasized the importance of ensuring the program’s sustainability. All the healthcare workers had a positive view of the program and acknowledged the strength of the program as portrayed by the excerpt below.

> *“The only thing I will like to say that it’s this a good program that have come to help these young ones”* (Key Informant Interview Participant, Healthcare Worker, LEKMA)

> *“The support group is open to all adolescents living with HIV because if I may be able to have my materials needs, I may be able to have my social needs, or emotional needs”* (Key Informant Interview Participant, Healthcare Worker, TEMA)

The healthcare workers acknowledged the program’s diverse merits, as illustrated by the extract.

> *“I think collaboration and coordination is key has been the key strength because… those linkage we have created is a game changer.”* (Key Informant Interview Participant, Healthcare Worker, CHAG)

> ” [The CATS program provides] *social support, more tailored care”* (Key Informant Interview Participant, Healthcare Worker, LEKMA)

> *“The strengths of the program are that the people are able to meet when they have to and gather to learn a lot about their condition and how best they can cope with it and how they can encourage others in similar situation.” (Key Informant Interview Participant, Healthcare Worker, TEMA)*

The program has achieved remarkable progress in empowering peer supporters and their clients, and their supervisors, as highlighted by the following excerpts.

> *“We have been able to support some of these CATS themselves to learn skills… we trained all the supervisor to lead support group session for the young ones in the facility.’* (Key Informant InterviewParticipant, Healthcare Worker, CHAG)

> *“…is building up confidence like public speech people are most of them are not able to be vocal.”* (Key Informant Interview Participant, Healthcare Worker, LEKMA)

The program nurtures an environment where the voices of the youth are amplified, their needs addressed, and a profound sense of inclusion and support is fostered. The principle of adolescent representation is a cornerstone of the program’s approach, as substantiated by the following excerpts.

> *“…in fact every single meeting that we have the young ones are duly represented in those meeting,so that we can also get their perspective as well’.* (Key Informant Interview Participant, Healthcare Worker, CHAG)

> *‘Initially, when they come for refills, they are in the back ground they weren’t even talking something it’s their parents were those coming forward, but since the group started, most of them are now coming on their own’* (Key Informant Interview Participant, Healthcare Worker, LEKMA)

The program places a strong emphasis on interaction and support, establishing a nurturing environment for its participants as indicated by the following.

> *“We organized support group meeting for them if they have any challenges there are able to discuss there and this has been a game changer.”* (Key Informant Interview Participant, Healthcare Worker, CHAG)

> *“…is that kind of friendship and relationship we have with them that helps us it’s not just me it’s all the staff.” (Key Informant Interview Participant, Healthcare Worker, LEKMA)*

> *‘I’m always available it’s a one stop place for them’.* (Key Informant Interview Participant,Healthcare Worker, TEMA)

While the program has made commendable strides, it also grapples with certain challenges that warrant careful consideration and concerted efforts towards resolution.

> *“It’s not easy to even recruit a CATS, their recruitment itself is …a major challenge.”* (Key Informant Interview Participant, Healthcare Worker, CHAG)

> *“Challenge so far so far the as I said support in terms of finances.”* (Key Informant Interview Participant, Healthcare Worker, TEMA)

In charting the way forward, healthcare workers have put forth valuable suggestions to make the program successful, as demonstrated by the following segment.

> *“Plan activities that we go out of the facility and find some where to go and if it is a beach side or the Pool side. You know, if there are activities, I’m sure a lot of them will want to come.”* (Key Informant Interview Participant, Healthcare Worker, LEKMA)

> *‘The people here supporting the clients also need to be motivated to get it because for some people it may not be their duty day but may need to come around because it’s a support group day”* (Key Informant Interview Participant, Healthcare Worker, TEMA)

### Discussion

#### Effectiveness of community adolescent treatment support in facilitating HIV care linkage and follow-up among adolescents

This study findings highlighted the Community Adolescent Treatment Support (CATS) program’s effectiveness in HIV care linkage and retention success by attracting clients, especially through engagement at standard ARV clinics, with a greater interest from girls. This is in line with the literature that emphasizes the importance of targeted outreach strategies in engaging individuals in HIV care[31,32]. The diverse motivations for joining the program underlined the need for personalized and different approaches to address the unique needs of adolescents living with HIV [9,33]. The study revealed that CATS functions as a vital support system, addressing emotional, social, and psychological needs of adolescents. The role of peer supporters in maintaining contact with clients and offering support beyond standard clinic hours echoed the importance of peer support programs in improving the well-being of adolescents living with HIV [6]. The provision of emotional and social support within the program aligns with the recommendation of offering comprehensive care to address the holistic needs of these individuals [34,35]. This present study findings suggest that the CATS program has significantly improved adherence and retention in care, resulting in better viral load outcomes and reduced hospital visits. These findings are consistent with existing research, which demonstrated that support programs can positively impact medication adherence and, consequently, health outcomes [12,36]. Improved adherence and continuity of care are crucial for adolescents living with HIV, as they contribute to better health outcomes and an improved quality of life [37].

On the other hand, this study identified challenges in CATS implementation, such as communication issues, financial barriers, and participant consistency. These challenges are not uncommon in healthcare programs, which underlined the importance of continuous improvement and adaptation [38,39]. Addressing communication issues through more efficient strategies, such as text messages or reminder calls, can enhance program effectiveness. Providing financial support could also improve program accessibility for those facing economic barriers. Strategies for accommodating participants’ schedules, such as offering flexible meeting times, can further help ensure consistent participation [40].

#### Acceptance of Community Adolescent Treatment Support by Adolescents

The findings demonstrated a high level of acceptance of the CATS program among adolescents living with HIV. This finding corroborated with the findings reported in a previous study conducted in Kumasi [41]. The adolescents valued the companionship and support provided by the support group. The acceptance of Mhealth services, group meetings, and home visit services underscored the importance of offering diverse services to cater to such individual preferences [9,42,43]. The positive reception of Mhealth services aligns with the growing role of technology in improving healthcare access and communication for adolescents [42,43]. The heartening enthusiasm of adolescents participating in the CATS program is remarkable. They found solace and a sense of community within group meetings, echoing the importance of peer support and shared experiences in adolescents’ living with HIV. Existing research has consistently emphasized the power of support groups in enhancing emotional well-being, adherence to treatment, and overall health outcomes among adolescents living with HIV [44,45]. This current study’s findings highlighted the need for interventions that create safe spaces for adolescents to share and relate to one another. The integration of mobile health (Mhealth) services is a standout feature of the CATS program. This approach leverages technology to facilitate communication and support between CATS and clients. The participants’ unanimous acceptance of Mhealth services demonstrates its potential to enhance accessibility and provide timely assistance, as reported in prior study [46]. However, challenges such as varying access to smartphones and concerns about privacy should be addressed. Strategies to offer more secure and inclusive Mhealth solutions may further improve program effectiveness [46].

This study identified the CATS program’s group meetings as a critical component, offering adolescents living with HIV a platform to share experiences, receive mutual support, and enhance their self-sufficiency. While most participants preferred support group meetings, some expressed discomfort among members. The findings echoed the importance of both group and individualized care approaches in addressing the diverse needs of adolescents with HIV as noted in previous studies [43, 45]. Home visit services play a crucial role in providing personalized care and support to adolescents. However, participants’ reluctance to embrace this service underscored the impact of stigma and privacy concerns. Prior studies have also noted the challenges related to home-based care due to issues of confidentiality and fear of disclosure [47].

#### Community factors that influence the implementation of Community AdolescentTreatment Support

The findings in this present study further highlighted the pervasive challenges of stigma and discrimination within the broader community. Adolescents living with HIV continue to face misunderstandings, fear, and isolation. This aligned with extensive literature that underscored the enduring stigma associated with HIV and its negative impact on individuals’ well-being [48]. It was also revealed that the participants in this study noted that many community members lack knowledge about HIV and hold misconceptions about its transmission. These findings supported what previous study has reported that community education and awareness programs were needed to dispel misconceptions and reduce HIV-related stigma [49]. This finding implied that adolescents perceive their communities as unsupportive. This lack of support can lead to feelings of isolation and reluctance to disclose their HIV status to others. Previous studies also emphasized the importance of community support networks for individuals living with HIV [50]. However, it was revealed that healthcare workers involved in the program are actively engaged in initiatives aimed at reducing stigma and discrimination. The involvement of healthcare workers in initiatives to fight stigma and discrimination is essential and resonates with broader efforts to create more supportive healthcare systems [51].

#### Stakeholder’s Perceptive on Community Adolescent Treatment Support

Healthcare professionals’ positive views of the CATS program are encouraging. Their acknowledgment of the program’s ability in addressing the distinctive psychosocial requirements of adolescents living with HIV, a group often neglected in the healthcare system. The positive feedback aligned with what is reported in existing literature emphasizing the importance of tailored support for adolescents living with HIV [52]. The program’s ability to amplify the voices of these adolescents is particularly noteworthy in this study. It was reported that the program empowers them to express their concerns and aspirations. This emphasis on empowering the youth aligns with the broader goals of adolescent-friendly healthcare services [53]. This implied that the idea of empowering adolescents in managing their health leads to better health outcomes [54]. The healthcare workers identified several key strengths of the CATS program, including collaboration and coordination, social support, and tailored care. Collaboration and coordination are essential for effective healthcare. The program’s focus on social support is crucial because it addresses the psychosocial needs of adolescents living with HIV [55]. The program’s commitment to amplifying the voices of young people living with HIV is also commendable. This approach recognizes the importance of involving adolescents in decisions about their care, which is a central principle of patient-centered care [56]. Involving adolescents in healthcare decision-making can improve adherence to treatment and overall well-being [56, 57].

### Conclusion

In conclusion, the Community Adolescent Treatment Support (CATS) program has demonstrated its effectiveness in improving the lives of adolescents living with HIV by providing a secured and supportive environment, enhancing medication adherence, and addressing the unique psychosocial needs of this population. While the program has made significant strides, challenges related to communication, financial barriers, and stigma persist. The acceptance of the CATS program among adolescents living with HIV is evident, and it offers a variety of services to cater to individual preferences. However, community factors, such as stigma and discrimination, continue to hinder social interactions and support.

Healthcare professionals recognize the program’s value in addressing the distinct needs of adolescents and emphasize collaboration and patient-centered care. Moving forward, it is crucial to address the identified challenges and enhance the program’s effectiveness to maximize its impact on the well-being of adolescents living with HIV, and community education and sensitization efforts are essential to create a more supportive and inclusive environment.

This study recommends that communication strategies, increasing funding support, and intensifying community education and sensitization efforts should be strengthened to reduce stigma and foster broader community inclusivity for adolescents living HIV and the CATS program.

**Table 1:** Socio-demographic characteristics of participants.

| **In-depth Interviews & Key Informant Interviews (N=16)** |  |  |
| --- | --- | --- |
| **Variables** | **Frequency(N=16)** | **Percentage (%)** |
| **Participant** |  |  |
| Clients | 10 | 62.5 |
| Peer Supporters | 3 | 18.8 |
| Healthcare Workers | 3 | 18.8 |
| **Sex** |  |  |
| Male | 6 | 37.5 |
| Female | 10 | 62.5 |
| **Age** |  |  |
| 18- 29 | 13 | 81.3 |
| 30-34 | 2 | 12.5 |
| 40 and above | 1 | 6.3 |
| **Educational Level** |  |  |
| No Formal Education | 2 | 12.5 |
| Basic | 5 | 31.3 |
| Secondary | 2 | 12.5 |
| Tertiary | 7 | 43.8 |
| **Employment Status** |  |  |
| Employed | 11 | 68.8 |
| Unemployed | 5 | 31.2 |
| **Marital Status** |  |  |
| Single | 10 | 62.5 |
| In a Relationship | 2 | 12.5 |
| Married | 4 | 25 |
| **Location** |  |  |
| CHAG | 3 | 18.8 |
| LEKMA | 7 | 43.8 |
| TEMA | 6 | 37.5 |

**Table 4.2.**
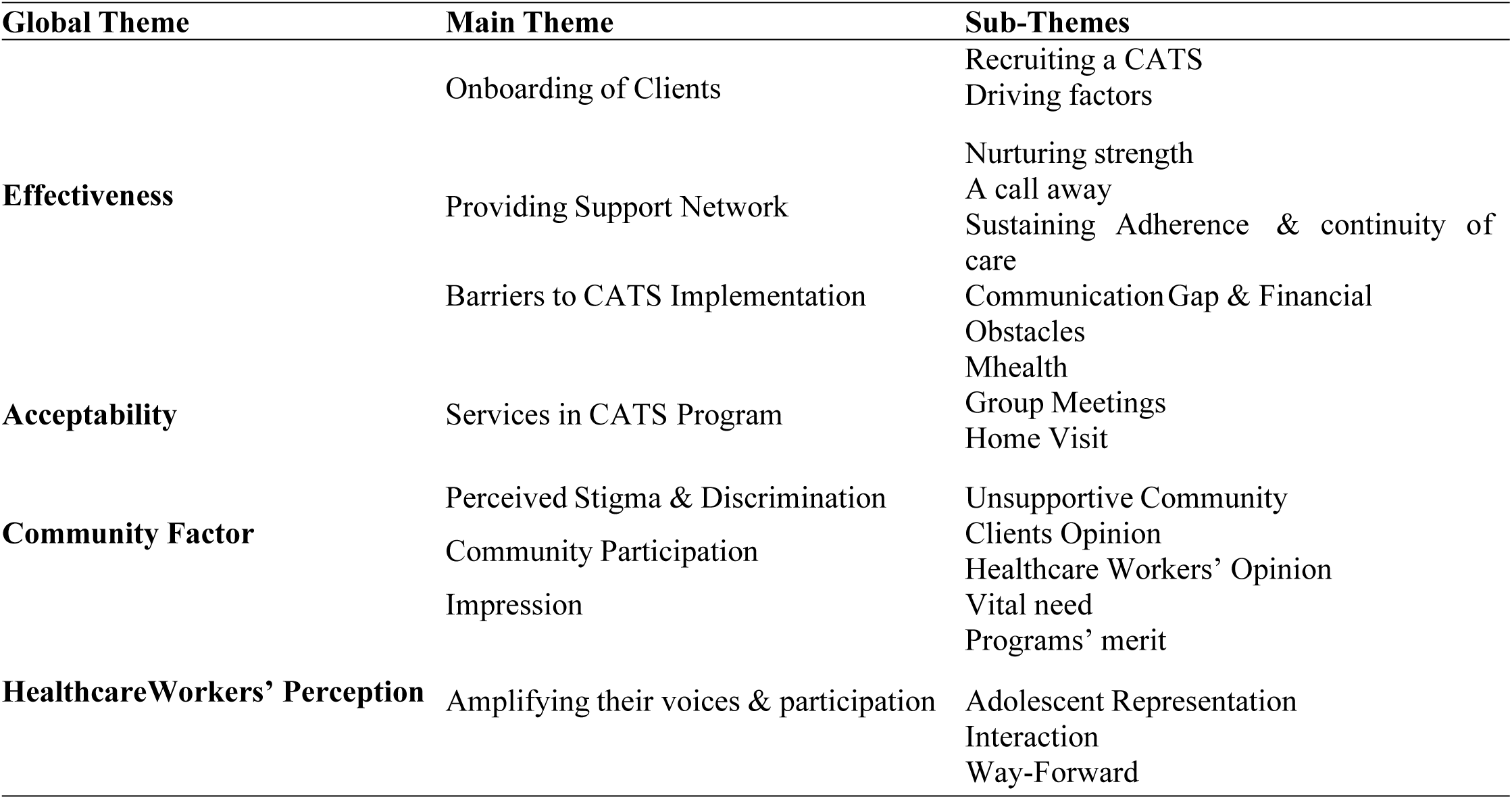
Themes That Emerged from The Data.

## Data Availability

The data for this study can be obtained from the corresponding author upon reasonable request through

## Acknowledgement

We are grateful to the Greater Accra Regional Health Directorate and various health facilities management for granting us permission to conduct this study. Many appreciations to the study participants for consenting to fully participate in this study.

## Notes

### Competing Interest Statement

The authors have declared no competing interest.

### Funding Statement

This study was funded by the Office of Research, Innovation, and Development under the WHO/TDR postgraduate training scheme.

### Author Declarations

Ethical clearance was sought from the Ghana Health Service Ethics Review Committee with reference number: GHS-ERC:070/05/23. Permission was sought from the Greater Accra Regional Health Directorate and from the management of the hospitals and the respective heads of department of each health facility. The participants signed a participant information sheet with an informed consent form before the data collection and interview. Participants were also allowed to decide not to participate, ask questions if they thought it was uncomfortable, or leave the study whenever they wanted. The discussion and interviews were conducted in a private setting for privacy purposes. All data received was protected and was not accessible to individuals who were not part of the team. Participants were made aware that the findings of this would be published in peer-reviewed journals. The findings and recommendations from this study were shared with the study participants and the hospitals where the study took place.

## Reference

[1] “UNAIDS-Inputs-to-the-Report-of-the-Secretary-General-2020”.

[2] I. J. K. Biney et al., “Antiretroviral therapy adherence and viral suppression among HIV-infected adolescents and young adults at a tertiary hospital in Ghana,” African Journal of AIDS Research, vol. 20, no. 4, pp. 270–276, 2021, doi: 10.2989/16085906.2021.1998783.

[3] O. A. Adejumo, K. M. Malee, P. Ryscavage, S. J. Hunter, and B. O. Taiwo, “Contemporary issues on the epidemiology and antiretroviral adherence of HIV-infected adolescents in sub-Saharan Africa: A narrative review,” J Int AIDS Soc, vol. 18, no. 1, Sep. 2015, doi: 10.7448/IAS.18.1.20049.

[4] L. Katirayi, J. Akuno, B. Kulukulu, and R. Masaba, “‘When you have a high life, and you like sex, you will be afraid’: a qualitative evaluation of adolescents’ decision to test for HIV in Zambia and Kenya using the health belief model,” BMC Public Health, vol. 21, no. 1, Dec. 2021, doi: 10.1186/s12889-021-10391-x.

[5] P. Nyakato et al., “Virologic non-suppression and early loss to follow up among pregnant and non-pregnant adolescents aged 15-19 years initiating antiretroviral therapy in South Africa: a retrospective cohort study,” J Int AIDS Soc, vol. 2022, p. 25870, 2022, doi: 10.1002/jia2.25870/full.

[6] T. Tapera et al., “Effects of a Peer-Led Intervention on HIV Care Continuum Outcomes Among Contacts of Children, Adolescents, and Young Adults LivingWith HIV in Zimbabwe,” Glob Health Sci Pract, vol. 7, no. 4, pp. 575–584, 2019, doi: 10.9745/GHSP-D-19-00210.

[7] K. Govender, W. G. B. Masebo, P. Nyamaruze, R. G. Cowden, B. T. Schunter, and A. Bains, “HIV Prevention in Adolescents and Young People in the Eastern and Southern African Region: A Review of Key Challenges Impeding Actions for an Effective Response,” Open AIDS J, vol. 12, no. 1, pp. 53–67, Jul. 2018, doi: 10.2174/1874613601812010053.

[8] “GHANA AIDS COMMISSION National and Sub-National HIV and AIDS Estimates and Projections 2017 Report.”

[9] S. Bernays et al., “Scaling up peer-led community-based differentiated support for adolescents living with HIV: keeping the needs of youth peer supporters in mind to sustain success,” J Int AIDS Soc, vol. 23, no. S5, pp. 15–20, 2020, doi: 10.1002/jia2.25570.

[10] N. Willis et al., “Effectiveness of community adolescent treatment supporters (CATS) interventions in improving linkage and retention in care, adherence to ART and psychosocial well-being: A randomised trial among adolescents living with HIV in rural Zimbabwe,” BMC Public Health, vol. 19, no. 1, pp. 1–9, 2019, doi: 10.1186/s12889-019-6447-4.

[11] P. Dako-Gyeke, B. Dornoo, S. Ayisi Addo, M. Atuahene, N. A. Addo, and A. E. Yawson, “Towards elimination of mother-to-child transmission of HIV in Ghana: An analysis of national programme data,” Int J Equity Health, vol. 15, no. 1, Jan. 2016, doi: 10.1186/s12939-016-0300-5.

[12] D. Barker et al., “In-Clinic Adolescent Peer Group Support for Engagement in Sub-Saharan Africa: A Feasibility and Acceptability Trial,” J Int Assoc Provid AIDS Care, vol. 18, Mar. 2019, doi: 10.1177/2325958219835786.

[13] A. Enimil et al., “Quality of life among Ghanaian adolescents living with perinatally acquired HIV: A mixed methods study,” AIDS Care - Psychological and Socio-Medical Aspects of AIDS/HIV, vol. 28, no. 4, pp. 460–464, Apr. 2016, doi: 10.1080/09540121.2015.1114997.

[14] E. O. Agyemang et al., “Self-Esteem Assessment among Adolescents Living with HIV and Seeking Healthcare at Komfo Anokye Teaching Hospital-Kumasi, Ghana,” J Int Assoc Provid AIDS Care, vol. 19, 2020, doi: 10.1177/2325958220976828.

[15] A. Brédart, A. Marrel, L. Abetz-Webb, K. Lasch, and C. Acquadro, “Interviewing to develop Patient-Reported Outcome (PRO) measures for clinical research: Eliciting patients’ experience,” Feb. 05, 2014. doi: 10.1186/1477-7525-12-15.

[16] H. Mohajan and H. K. Mohajan, “Munich Personal RePEc Archive Qualitative Research Methodology in Social Sciences and Related Subjects Qualitative Research Methodology in Social Sciences and Related Subjects,” 2018.

[17] I. Etikan, “Comparison of Convenience Sampling and Purposive Sampling,” American Journal of Theoretical and Applied Statistics, vol. 5, no. 1, p. 1, 2016, doi: 10.11648/j.ajtas.20160501.11.

[18] G. Guest, A. Bunce, and L. Johnson, “How Many Interviews Are Enough?: An Experiment with Data Saturation and Variability,” Field methods, vol. 18, no. 1, pp. 59–82, 2006, doi: 10.1177/1525822X05279903.

[19] F. T. Gyimah and P. Dako-Gyeke, “Perspectives on TB patients’ care and support: A qualitative study conducted in Accra Metropolis, Ghana,” Global Health, vol. 15, no. 1, Mar. 2019, doi: 10.1186/s12992-019-0459-9.

[20] M. Cleary, J. Horsfall, and M. Hayter, “Data collection and sampling in qualitative research: Does size matter?,” Mar. 2014. doi: 10.1111/jan.12163.

[21] M. M. Hennink, B. N. Kaiser, and M. B. Weber, “What Influences Saturation? Estimating Sample Sizes in Focus Group Research,” Qual Health Res, vol. 29, no. 10, pp. 1483–1496, Aug. 2019, doi: 10.1177/1049732318821692.

[22] P. I. Fusch and L. R. Ness, “Are we there yet? Data saturation in qualitative research,” Qualitative Report, vol. 20, no. 9, pp. 1408–1416, Sep. 2015, doi: 10.46743/2160-3715/2015.2281.

[23] L. A. Palinkas, G. A. Aarons, S. Horwitz, P. Chamberlain, M. Hurlburt, and J. Landsverk, “Mixed method designs in implementation research,” Administration and Policy in Mental Health and Mental Health Services Research, vol. 38, no. 1, pp. 44–53, Jan. 2011, doi: 10.1007/s10488-010-0314-z.

[24] L. A. Palinkas, G. A. Aarons, S. Horwitz, P. Chamberlain, M. Hurlburt, and J. Landsverk, “Mixed method designs in implementation research,” Administration and Policy in Mental Health and Mental Health Services Research, vol. 38, no. 1, pp. 44–53, Jan. 2011, doi: 10.1007/s10488-010-0314-z.

[25] E. Knott, A. H. Rao, K. Summers, and C. Teeger, “Interviews in the social sciences,” Nature Reviews Methods Primers, vol. 2, no. 1, Dec. 2022, doi: 10.1038/s43586-022-00150-6.

[26] J. M. Morse, “Critical Analysis of Strategies for Determining Rigor in Qualitative Inquiry,” in Qualitative Health Research, SAGE Publications Inc., Sep. 2015, pp. 1212–1222. doi: 10.1177/1049732315588501.

[27] A. Moser and I. Korstjens, “Series: Practical guidance to qualitative research. Part 3: Sampling, data collection and analysis,” Jan. 01, 2018, Taylor and Francis Ltd. doi: 10.1080/13814788.2017.1375091.

[28] V. Braun and V. Clarke, “Using thematic analysis in psychology,” Qual Res Psychol, vol. 3, no. 2, pp. 77–101, 2006, doi: 10.1191/1478088706qp063oa.

[29] J. Fereday, N. Adelaide, S. Australia, and A. Eimear Muir-Cochrane, “Demonstrating Rigor Using Thematic Analysis: A Hybrid Approach of Inductive and Deductive Coding and Theme Development,” 2006.

[30] M. Arain et al., “Maturation of the adolescent brain,” 2013, Dove Medical Press Ltd. doi: 10.2147/NDT.S39776.

[31] P. Dako-Gyeke, R. Snow, and A. E. Yawson, “Who is utilizing anti-retroviral therapy in Ghana: An analysis of ART service utilization,” Int J Equity Health, vol. 11, no. 1, p. 62, 2012, doi: 10.1186/1475-9276-11-62.

[32] W. Mavhu et al., “Enhancing Psychosocial Support for HIV Positive Adolescents in Harare, Zimbabwe,” PLoS One, vol. 8, no. 7, Jul. 2013, doi: 10.1371/journal.pone.0070254.

[33] J. A. Denison et al., “The sky is the limit: Adhering to antiretroviral therapy and HIV self-management from the perspectives of adolescents living with HIV and their adult caregivers,” J Int AIDS Soc, vol. 18, no. 1, Jan. 2015, doi: 10.7448/IAS.18.1.19358.

[34] D. Baron et al., “‘you talk about problems until you feel free’: South African adolescent girls’ and young women’s narratives on the value of HIV prevention peer support clubs,” BMC Public Health, vol. 20, no. 1, Jun. 2020, doi: 10.1186/s12889-020-09115-4.

[35] L. Cluver, M. Pantelic, M. Orkin, E. Toska, S. Medley, and L. Sherr, “Sustainable Survival for adolescents living with HIV: do SDG-aligned provisions reduce potential mortality risk?,” 2018, doi: 10.1002/jia2.25056/full.

[36] J. Johnson-Peretz et al., “‘I was still very young’: agency, stigma and HIV care strategies at school, baseline results of a qualitative study among youth in rural Kenya and Uganda,” J Int AIDS Soc, vol. 2022, no. S1, p. 25919, 2022, doi: 10.1002/jia2.25919/full.

[37] C. Nimwesiga, I. M. Taremwa, D. Nakanjako, and E. Nasuuna, “Factors Associated with Retention in HIV Care Among HIV-Positive Adolescents in Public Antiretroviral Therapy Clinics in Ibanda District, Rural South Western Uganda,” HIV/AIDS - Research and Palliative Care, vol. 15, pp. 71–81, 2023, doi: 10.2147/HIV.S401611.

[38] L. A. Enane et al., “‘A problem shared is half solved’–a qualitative assessment of barriers and facilitators to adolescent retention in HIV care in western Kenya,” AIDS Care - Psychological and Socio-Medical Aspects of AIDS/HIV, vol. 32, no. 1, pp. 104–112, Jan. 2020, doi: 10.1080/09540121.2019.1668530.

[39] E. C. Kip, M. Udedi, K. Kulisewa, V. F. Go, and B. N. Gaynes, “Stigma and mental health challenges among adolescents living with HIV in selected adolescent-specific antiretroviral therapy clinics in Zomba District, Malawi,” BMC Pediatr, vol. 22, no. 1, pp. 1–12, 2022, doi: 10.1186/s12887-022-03292-4.

[40] L. K. Beres et al., “Trajectories of re-engagement: factors and mechanisms enabling patient return to HIV care in Zambia,” J Int AIDS Soc, vol. 26, p. 26067, 2023, doi: 10.1002/jia2.26067/full.

[41] D. Barker et al., “In-Clinic Adolescent Peer Group Support for Engagement in Sub-Saharan Africa: A Feasibility and Acceptability Trial,” J Int Assoc Provid AIDS Care, vol. 18, Mar. 2019, doi: 10.1177/2325958219835786.

[42] V. Bertman et al., “Health worker text messaging for blended learning, peer support, and mentoring in pediatric and adolescent HIV/AIDS care: A case study in Zimbabwe,” Hum Resour Health, vol. 17, no. 1, Jun. 2019, doi: 10.1186/s12960-019-0364-6.

[43] E. Toska et al., “Screening and supporting through schools: Educational experiences and needs of adolescents living with HIV in a South African cohort,” BMC Public Health, vol. 19, no. 1, Mar. 2019, doi: 10.1186/s12889-019-6580-0.

[44] E. A. Abrams et al., “Adolescents do not only require arvs and adherence counseling“: A qualitative investigation of health care provider experiences with an HIV youth peer mentoring program in ndola, Zambia,” PLoS One, vol. 16, no. 6 June, Jun. 2021, doi: 10.1371/journal.pone.0252349.

[45] D. Mark et al., “Peer Support for Adolescents and Young People Living with HIV in sub-Saharan Africa: Emerging Insights and a Methodological Agenda,” Dec. 01, 2019, Springer. doi: 10.1007/s11904-019-00470-5.

[46] A. Haleem, M. Javaid, R. P. Singh, and R. Suman, “Telemedicine for healthcare: Capabilities, features, barriers, and applications,” Jan. 01, 2021, KeAi Communications Co. doi: 10.1016/j.sintl.2021.100117.

[47] J. S. Ambikile and M. K. Iseselo, “Challenges to the provision of home care and support for people with severe mental illness: Experiences and perspectives of patients, caregivers, and healthcare providers in Dar es Salaam, Tanzania,” PLOS Global Public Health, vol. 3, no. 1, p. e0001518, Jan. 2023, doi: 10.1371/journal.pgph.0001518.

[48] E. Kimera et al., “Experiences and effects of HIV-related stigma among youth living with HIV/AIDS in Western Uganda: A photovoice study,” PLoS One, vol. 15, no. 4, Apr. 2020, doi: 10.1371/journal.pone.0232359.

[49] I. C. Agu et al., “Misconceptions about transmission, symptoms and prevention of HIV/AIDS among adolescents in Ebonyi state, South-east Nigeria,” BMC Res Notes, vol. 13, no. 1, May 2020, doi: 10.1186/s13104-020-05086-2.

[50] L. Aurpibul et al., “Social effects of HIV disclosure, an ongoing challenge in young adults living with perinatal HIV: a qualitative study,” Front Public Health, vol. 11, 2023, doi: 10.3389/fpubh.2023.1150419.

[51] L. Nyblade et al., “Stigma in health facilities: Why it matters and how we can change it,” BMC Med, vol. 17, no. 1, Feb. 2019, doi: 10.1186/s12916-019-1256-2.

[52] J. K. B. Matovu, A. Nambuusi, S. Nakabirye, R. K. Wanyenze, and D. Serwadda, “Formative research to inform the development of a peer-led HIV self-testing intervention to improve HIV testing uptake and linkage to HIV care among adolescents, young people and adult men in Kasensero fishing community, Rakai, Uganda: a qualitative study,” BMC Public Health, vol. 20, no. 1, Dec. 2020, doi: 10.1186/s12889-020-09714-1.

[53] “ADOLESCENT-FRIENDLY HEALTH SERVICES FOR ADOLESCENTS LIVING WITH HIV: FROM THEORY TO PRACTICE TECHNICAL BRIEF PEER DRIVEN ADOLESCENT HIV MODELS OF CARE,” 2019. [Online]. Available: http://apps.who.int/bookorders.

[54] G. Antelman et al., “Adolescent support club attendance and self-efficacy associated with HIV treatment outcomes in Tanzania,” PLOS Global Public Health, vol. 2, no. 10, p. e0000065, Oct. 2022, doi: 10.1371/journal.pgph.0000065.

[55] B. N. Kaunda-Khangamwa et al., “Adolescents living with HIV, complex needs and resilience in Blantyre, Malawi,” AIDS Res Ther, vol. 17, no. 1, Jun. 2020, doi: 10.1186/s12981-020-00292-1.

[56] D. Mark et al., “HIV treatment and care services for adolescents: A situational analysis of 218 facilities in 23 sub-Saharan African countries,” J Int AIDS Soc, vol. 20, May 2017, doi: 10.7448/IAS.20.4.21591.

[57] S. A. Fleary and P. Joseph, “Adolescents’ Health Literacy and Decision-making: A Qualitative Study,” Am J Health Behav, vol. 44, no. 4, pp. 392–408, Jul. 2020, doi: 10.5993/AJHB.44.4.3.

